# Elapsed time since BNT162b2 vaccine and risk of SARS-CoV-2 infection in a large cohort

**DOI:** 10.1101/2021.08.03.21261496

**Authors:** Ariel Israel, Eugene Merzon, Alejandro A. Schäffer, Yotam Shenhar, Ilan Green, Avivit Golan-Cohen, Eytan Ruppin, Eli Magen, Shlomo Vinker

## Abstract

**Importance:** Israel was among the first countries to launch a large-scale COVID-19 vaccination campaign, and quickly vaccinated its population, achieving early control over the spread of the virus. However, the number of COVID-19 cases is now rapidly increasing, which may indicate that vaccine protection decreases over time.

**Objective:** To determine whether time elapsed since the second BNT162b2 messenger RNA (mRNA) vaccine (Pfizer-BioNTech) injection is significantly associated with the risk of post-vaccination COVID-19 infection.

**Design:** This is a retrospective cohort study performed in a large state-mandated health care organization in Israel.

**Participants:** All fully vaccinated adults who have received a RT-PCR test between May 15, 2021 and July 26, 2021, at least two weeks after their second vaccine injection were included. Patients with a history of past COVID-19 infection were excluded.

**Main Outcome and Measure:** Positive result for the RT-PCR test.

**Results:** The cohort included 33,993 fully vaccinated adults, 49% women, with a mean age of 47 years (SD, 17 years), who received an RT-PCR test for SARS-CoV-2 during the study period. The median time between the second dose of the vaccine and the RT-PCR test was 146 days, interquartile range [121-167] days. 608 (1.8%) patients had positive test results. There was a significantly higher rate of positive results among patients who received their second vaccine dose at least 146 days before the RT-PCR test compared to patients who have received their vaccine less than 146 days before: odds ratio for infection was 3.00 for patients aged over 60 (95% CI 1.86-5.11); 2.29 for patients aged between 40 and 59 (95% CI 1.67-3.17); and 1.74 for patients aged between 18 and 39 (95% CI 1.27-2.37); P<0.001 in each age group.

**Conclusions and Relevance:** In this large population study of patients tested for SARS-CoV-2 by RT-PCR following two doses of mRNA BNT162b2 vaccine, we observe a significant increase of the risk of infection in individuals who received their last vaccine dose since at least 146 days ago, particularly among patients older than 60.

## Introduction

Immunity to severe acute respiratory syndrome coronavirus 2 (SARS-CoV-2) has been induced either through SARS-CoV-2 infection or vaccination and induces protection against reinfection or decreases the risk of clinically significant consequences^1^. While convalesced seropositive individuals have approximately 90% protection from SARS-CoV-2 reinfection, the effectiveness of vaccination has been reported as 50 - 95%^2–4^. Nevertheless, both the memory B cell humoral response and spike-specific CD4^+^ cellular immune responses to SARS-CoV-2 are predictably diminishing over time^5,6^. Therefore, there is a great concern regarding possible weakening of SARS-CoV-2 immune protection both in the vaccinated and convalescent population^7^.

Israel was among the first countries to initiate a large-scale vaccination campaign, on December 20, 2020, and quickly reached a high rate of immunized population, achieving early control over the spread of the virus^8^. More than 5.2 million Israelis have been fully vaccinated by two doses of the Pfizer-BioNTech vaccine as of July 26, 2021^9^. However, since June 2021, there has been a resurgence of SARS-CoV-2 cases, which could be at least partially attributed to decreasing levels of anti-SARS-CoV-2 antibodies in vaccinated individuals^10^.

Here we describe the results of a large-scale study measuring the association between time elapsed since the administration of second doses of BNT162b2 vaccine and the risk for COVID-19 disease.

## Methods

The study protocol was approved by the statutory clinical research committee of Leumit Health Services and the Shamir Medical Center Institutional Review Board (129-2-LEU). Informed consent was waived because only deidentified routinely collected data were used.

### Study Population

This is a population-based study based on data from Leumit Health Services (LHS), a large nation-wide healthcare provider in Israel, which provides services to around 700,000 members throughout the country. LHS uses centrally managed electronic health records (EHRs), continuously updated regarding subjects’ demographics, medical diagnoses, medical encounters, hospitalizations and laboratory tests. All LHS members have similar health insurance and similar access to healthcare services.

All LHS members who have been fully vaccinated and underwent a SARS-CoV-2 PCR test between May 15, 2021 and July 26, 2021, with an additional timing criterion that the test was performed at least two weeks following their second vaccine injection, were included. Tests from individuals who were diagnosed with COVID-19 prior to the PCR test were excluded. Patients who requested more than one PCR test during the period were included only once, at the date of their last eligible test. We partitioned the cohort in three age groups (60 or above, between 40 and 59, and between 18 and 39 years), to reflect the vaccine rollout stages.

Baseline data regarding the cohort was extracted as of May 15, 2021, including age. All the clinical diagnoses were based on ICD-9 codes. We tested for the main medical conditions expected to affect the rates of COVID-19 infection in adult population: diabetes mellitus, hypertension, asthma, chronic obstructive pulmonary disease, ischemic heart disease, presence of malignancy, and chronic kidney disease. During each physician visit, a diagnosis is entered or updated according to the International Classification of Diseases 9^th^ revision (ICD-9). The validity of chronic diagnoses in the registry has been previously examined and confirmed as high^11,12^.

The Socio-economic status (SES) was defined according to a person’s home address. The Israeli Central Bureau of Statistics classifies all cities and settlements into 20 levels of SES. Ethnicity was also defined according to the home address of the HMO member, and categorized into three groups: General population, Ultra-Orthodox Jews and Arabs.

### SARS-CoV-2 testing by real-time RT-PCR

Nasopharyngeal swabs were taken and examined for SARS-CoV-2 by real-time RT-PCR performed with internal positive and negative controls, according to World Health Organization guidelines. The COBAS SARS-Cov-2 6800/8800 assay (Roche Pharmaceuticals, Basel, Switzerland) has been used since March 10, 2020.

### Study Outcomes

The primary outcome was SARS-CoV-2 infection detected by the RT-PCR test.

### Statistical Analyses

Standard descriptive statistics were used to present the demographic characteristics of individuals included in the study. The two-sided Wilcoxon rank sum test was used to compare continuous variables and Fisher’s test to compare categorial variables. Time elapsed since the second dose of the vaccine and until RT-PCR test was compared between individuals who received positive vs. negative results. Choosing the median time of 146 days as the cut-off, we further compared the rate of positive results among patients who received their second vaccine dose since at least vs. less than the median time. Odds ratio were obtained using Fisher’s exact test. Adjusted odds ratio were obtained by fitting multivariable logistic regression models, using sex, SES, ethnic group, and comorbidity factors (diabetes mellitus, hypertension, asthma, chronic obstructive pulmonary disease, ischemic heart disease, presence of malignancy, and chronic kidney disease) as covariates; regression model for the whole cohort was also adjusted for age category. Percentages of positive results in the two groups are presented graphically, using standard error bars, with stars displaying the significance level.

A two sided P<0.5 was considered statistically significant.

Statistical analysis was performed with R version 4.0.2 (R Foundation for Statistical Computing).

## Results

A total of 85,346 individuals underwent RT-PCR tests for SARS-CoV-2 during the study period, of which 57,361 were adult patients aged between 18 and 99. Figure 1 displays the flow diagram used for cohort selection: 33,993 fully vaccinated adults, who received an RT-PCR test for SARS-CoV-2 during the study period, at least two weeks after their second vaccine injection, and with no evidence of previous COVID-19 infection, were included in the cohort.

**Figure 1:**
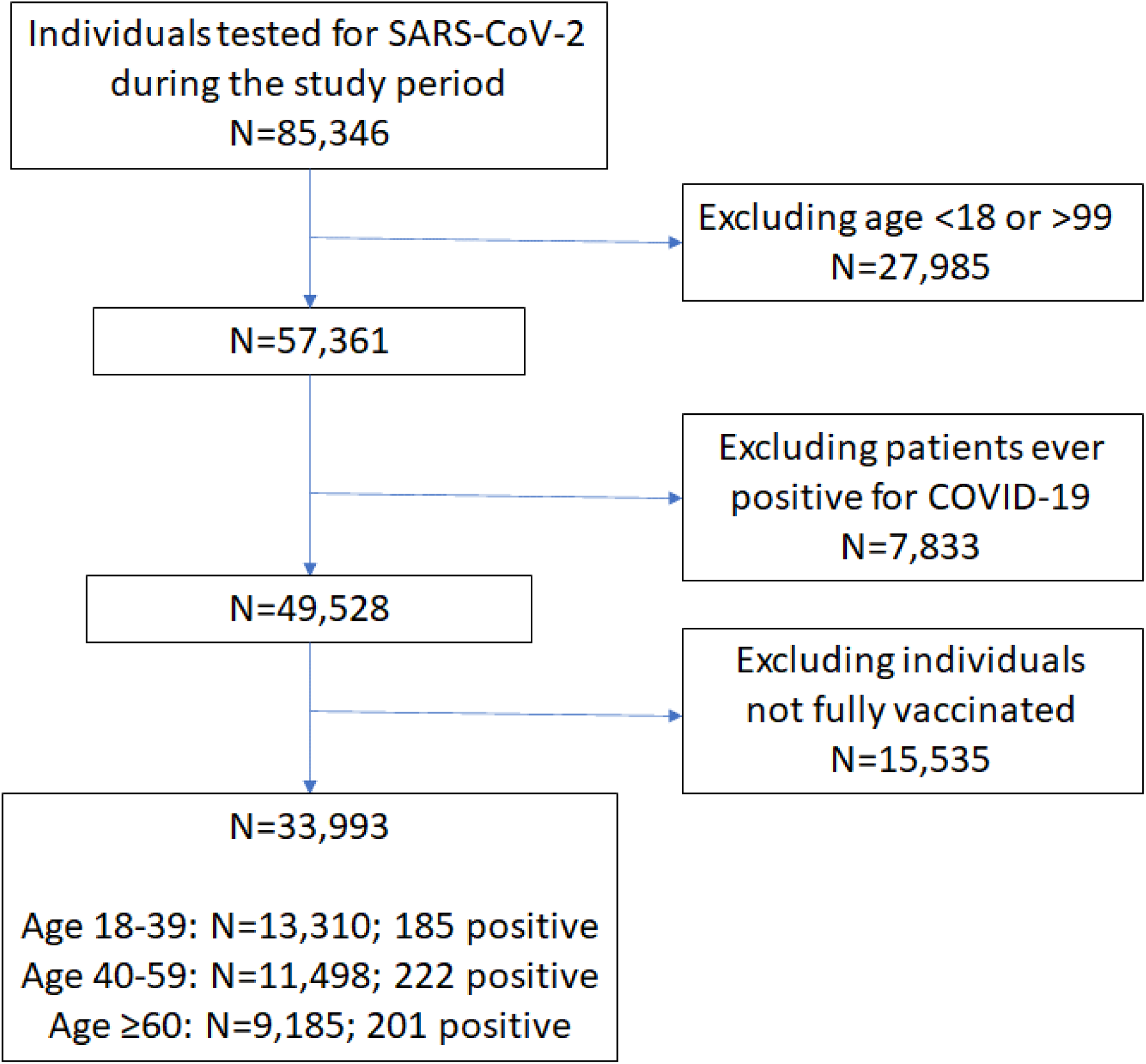
Flow diagram of the cohort.

**Figure 2:**
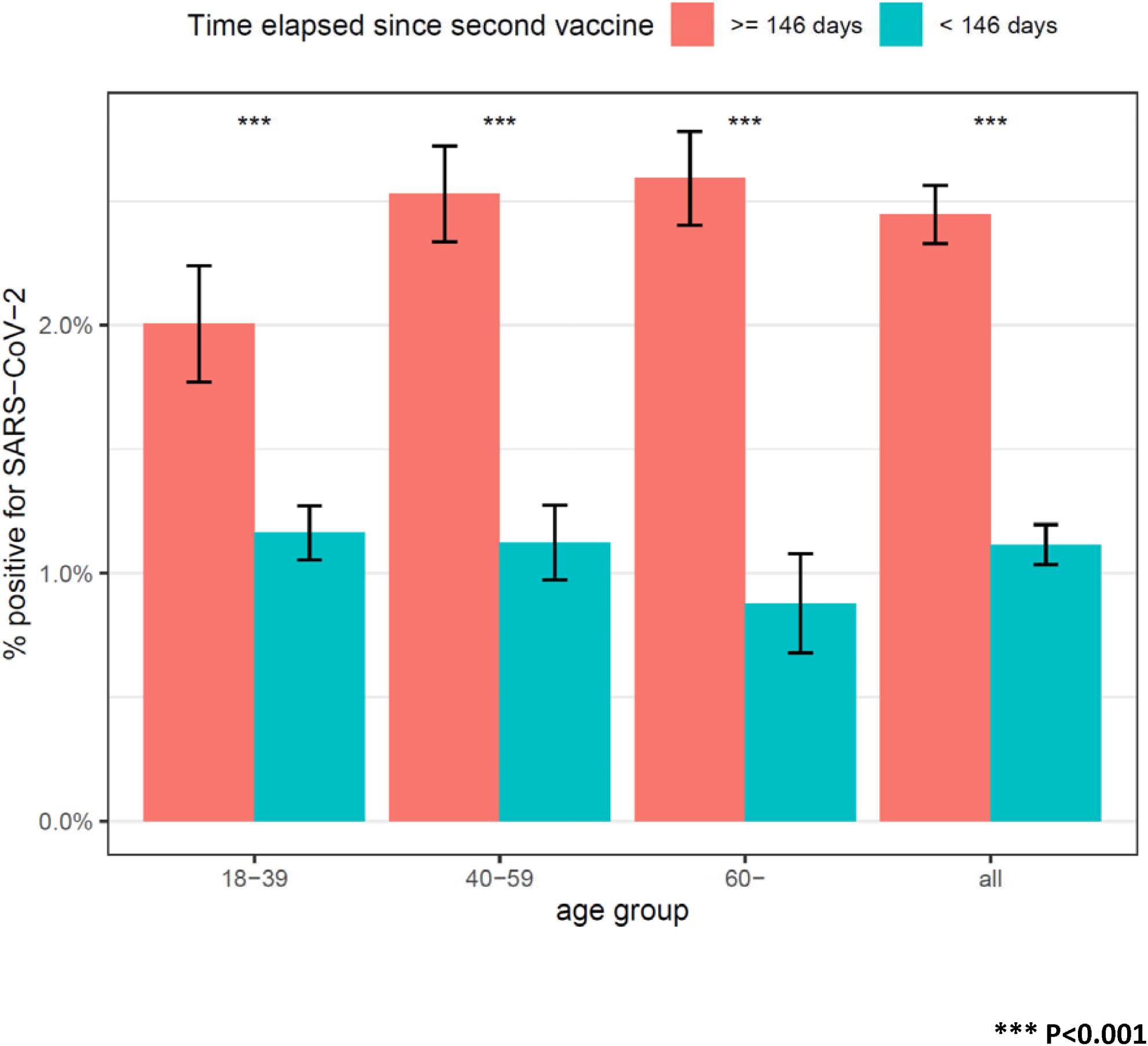
Comparison of the percentage of positive results among fully vaccinated individuals, according to time elapsed since the second vaccine dose.

The characteristics of the cohort are displayed in Table 1. Mean age is 46.8 years (SD, 17.4 years). 48.9% were female. 608 (1.8%) had positive test results. The median time between the second dose of the vaccine and the RT-PCR test was 146 days interquartile range (IQR) [121-167] days. Table 2 compares the characteristics of the patients who obtained a positive result vs. patients who tested negative. In each age group, the most significant difference between patients who tested positive and patients who tested negative was elapsed time (in days) since the date of the second dose of the vaccine. There was no significant difference in the interval between the first and second dose of the vaccine (almost all patients in our health organization received the second dose of the vaccine 21 days after the first); there were also no statistically significant differences in terms of baseline comorbidity factors, and except in the youngest age group, there were no significant differences in the age of patients who tested positive vs. patients tested negative. Statistically significant, but minor differences appeared in the SES of patients who tested positive in the younger age groups.

**Table 1:** Demographic and clinical characteristics of patients who performed a RT-PCR test during the study period and were included in the cohort.

|  |  |  |
| --- | --- | --- |
| **N** |  | 33,993 |
| **positive for SARS-CoV-2, N (%)** |  | 46.75 (17.41) |
| **age** | mean (SD) | 16,630 (48.9 %) |
|  | 18-39 | 13,310 (39.2 %) |
|  | 40-59 | 11,498 (33.8 %) |
|  | ≥ 60 | 9,185 (27.0 %) |
| **sex** | Female | 16,630 (48.9 %) |
|  | Male | 17,363 (51.1 %) |
| **socio-economic status (1-20)** | mean (SD) | 9.98 (3.74) |
|  | missing (%) | 3,154 (10.23 %) |
| **Ethnic group** | Arab | 5,143 (15.1 %) |
|  | General | 25,826 (76.0 %) |
|  | Jewish Ultra-orthodox | 3,024 (8.9 %) |
| **days elapsed** | since 2<sup>nd</sup> dose of the vaccine, median [IQR] | 146 [121-167] |
|  | ≥146 days since vaccination, N (%) | 17,211 (50.6 %) |
|  | between 1<sup>st</sup> and 2<sup>nd</sup> dose of vaccine, median [IQR] | 21 [21-21] |
| **comorbidities** | diabetes mellitus, N (%) | 4,351 (12.8 %) |
|  | hypertension, N (%) | 8,095 (23.8 %) |
|  | asthma, N (%) | 2,996 (8.8 %) |
|  | COPD, N (%) | 1,618 (4.8 %) |
|  | ischemic heart disease, N (%) | 2,038 (6.0 %) |
|  | solid tumor, N (%) | 2,085 (6.1 %) |
|  | chronic kidney disease, N (%) | 750 (2.2 %) |

**Table 2:** comparative analysis of individuals according to their RT-PCR result during the study period.

|  | Age ≥ 60 |  |  |  | age 40-59 |  |  |  | age 18-39 |  |  |  |
| --- | --- | --- | --- | --- | --- | --- | --- | --- | --- | --- | --- | --- |
|  | negative | positive | p val. | OR | negative | positive | p val. | OR | negative | positive | p val. | OR |
| **N** | 8,984 | 201 |  |  | 11,276 | 222 |  |  | 13,125 | 185 |  |  |
| **age, mean (SD)** | 69.24 (7.32) | 70.25 (7.86) | 0.055 |  | 49.36 (5.74) | 48.75 (5.58) | 0.117 |  | 28.97 (6.11) | 30.03 (6.12) | 0.019 |  |
| **sex female, N (%)** | 4,645 (51.7 %) | 95 (47.3 %) | 0.225 | 0.84 | 5,620 (49.8 %) | 115 (51.8 %) | 0.588 | 1.08 | 6,062 (46.2 %) | 93 (50.3 %) | 0.299 | 1.18 |
| **Socio-economic status, mean (SD)** | 10.94 (3.44) | 11.21 (3.23) | 0.274 |  | 10.16 (3.70) | 10.77 (3.48) | 0.021 |  | 9.11 (3.80) | 10.16 (3.45) | <0.001 |  |
| **time since 2<sup>nd</sup> dose of vaccine in days, median [IQR]** | 167 [147-179] | 175 [165-182] | <0.001 |  | 151 [128-166] | 159 [146-170] | <0.001 |  | 128 [105-147] | 141 [128-153] | <0.001 |  |
| **more than 146 days since vaccination, N (%)** | 6,839 (76.1 %) | 182 (90.5 %) | <0.001 | **3.00** | 6,433 (57.1 %) | 167 (75.2 %) | <0.001 | **2.29** | 3,518 (26.8 %) | 72 (38.9 %) | <0.001 | **1.74** |
| **time between 1<sup>st</sup> and 2<sup>nd</sup> dose of vaccine in days, median [IQR]** | 21 [21-21] | 21 [21-21] | 0.190 |  | 21 [21-21] | 21 [21-21] | 0.575 |  | 21 [21-21] | 21 [21-21] | 0.123 |  |
| **diabetes mellitus, N (%)** | 2,927 (32.6 %) | 72 (35.8 %) | 0.361 | 1.15 | 1,166 (10.3 %) | 20 (9.0 %) | 0.579 | 0.86 | 162 (1.2 %) | 4 (2.2 %) | 0.297 | 1.77 |
| **hypertension, N (%)** | 5,517 (61.4 %) | 131 (65.2 %) | 0.305 | 1.18 | 2,092 (18.6 %) | 35 (15.8 %) | 0.337 | 0.82 | 313 (2.4 %) | 7 (3.8 %) | 0.219 | 1.61 |
| **asthma, N (%)** | 865 (9.6 %) | 21 (10.4 %) | 0.716 | 1.10 | 852 (7.6 %) | 16 (7.2 %) | 1.000 | 0.95 | 1,226 (9.3 %) | 16 (8.6 %) | 0.898 | 0.92 |
| **COPD, N (%)** | 1,093 (12.2 %) | 27 (13.4 %) | 0.585 | 1.12 | 404 (3.6 %) | 5 (2.3 %) | 0.361 | 0.62 | 88 (0.7 %) | 1 (0.5 %) | 1.000 | 0.81 |
| **ischemic heart disease, N (%)** | 1,631 (18.2 %) | 37 (18.4 %) | 0.926 | 1.02 | 352 (3.1 %) | 6 (2.7 %) | 1.000 | 0.86 | 11 (0.1 %) | 1 (0.5 %) | 0.155 | 6.48 |
| **solid tumor, N (%)** | 1,446 (16.1 %) | 28 (13.9 %) | 0.496 | 0.84 | 486 (4.3 %) | 6 (2.7 %) | 0.313 | 0.62 | 116 (0.9 %) | 3 (1.6 %) | 0.230 | 1.85 |
| **chronic kidney disease, N (%)** | 602 (6.7 %) | 20 (10.0 %) | 0.086 | 1.54 | 91 (0.8 %) | 1 (0.5 %) | 1.000 | 0.56 | 34 (0.3 %) | 2 (1.1 %) | 0.089 | 4.21 |

OR: odds ratio; SD: Standard Deviation; IQR: Interquartile range; COPD: Chronic Obstructive Pulmonary Disease

Using the median of 146 days as a cut-off, we compared the positive rate in individuals who had received their second vaccine dose since at least 146 days vs. patients vaccinated earlier. Table 3 displays the odds ratio (OR) and 95% confidence intervals for testing positive among patients with at least 146 days of elapsed time since the second vaccine dose. The comparison is highly statistically significant (P<0.001) in all age groups: the odds ratio is highest for patients aged over 60, OR=3.00 (95% CI 1.86-5.12); but risk is also significantly increased for patients aged between 40 and 59 OR=2.29 (95% CI 1.67-3.17); and for patients aged between 18 and 39 OR=1.74 (95% CI 1.27-2.37); overall, the odds ratio for infection when vaccine was performed at least 146 days ago was 2.22. The adjusted odds ratio, obtained by fitting multivariable logistic regression models were also highly significant.

**Table 3:** odds ratio calculation for the risk of SARS-CoV-2 according to the time elapsed since the second SARS-CoV-2 (greater or lower than 146 days)

| age group<br>(years) | PCR test performed |  |  |  |  |  | Odds Ratio [CI] | P<br>value | Adjusted<br>odds Ratio [CI] * |
| --- | --- | --- | --- | --- | --- | --- | --- | --- | --- |
|  | ≥ 146 days since vaccine |  |  | < 146 days since vaccine |  |  |  |  |  |
|  | nb<br>patients<br>positive | nb<br>patients<br>tested | %<br>positive | nb<br>patients<br>positive | nb<br>patients<br>tested | %<br>positive |  |  |  |
| age 18-39 | 72 | 3,590 | 2.0% | 113 | 9,720 | 1.2% | 1.740 [1.273,2.365] | <0.001 | 1.67 [1.21,2.29] |
| age 40-59 | 167 | 6,600 | 2.5% | 55 | 4,898 | 1.1% | 2.286 [1.672,3.167] | <0.001 | 2.22 [1.62,3.08] |
| age ≥ 60 | 182 | 7,021 | 2.6% | 19 | 2,164 | 0.9% | 3.004 [1.863,5.118] | <0.001 | 2.76 [1.62-3.08] |
| all | 421 | 17,211 | 2.4% | 187 | 16,782 | 1.1% | 2.225 [1.866,2.661] | <0.001 | 2.06 [1.69,2.51] |

\* Adjusted odds ratio are adjusted for age group, sex, socio-economic status (SES), ethnic group, and comorbidity factors (diabetes mellitus, hypertension, asthma, chronic obstructive pulmonary disease, ischemic heart disease, presence of malignancy, and chronic kidney disease) as covariates.

## Discussion

In this large population of individuals who have received two doses of the BNT162b2 vaccine, we observe a significantly higher risk of SARS-CoV-2 infection among patients who have received their second vaccine dose since at least 146 days. The increase was significant for all age groups, with the strongest increase observed for patients aged 60 or more, with an odds ratio for infection of 3.00. These results are consistent with the general concept that the immune response to vaccines is influenced by age-related changes within the immune system^13^. This study likewise supports the findings reported by Lumley et al. ^2^, but the cohort in this study is larger and more generalizable to the general population.

The strengths of this analysis include the use of a large cohort of fully vaccinated individuals, all of whom have received the same vaccine, with detailed demographic and clinical information and continuously updated data on vaccination and SARS-CoV-2 infection status. Israel was one of the first countries to rollout a large-scale vaccination campaign, so patients included in this study have received their second vaccine injection up to 6 months ago. To our knowledge, this is the first study to provide clear evidence of a decreasing COVID-19 vaccine protection with increasing time from the second injection. Throughout the study period, most of the new infections by SARS-CoV-2 were with the delta variant B.1.617.2 (93% of 113 isolates sent for sequencing in our health organization), so our study reflects the protection offered by the vaccine against the SARS-CoV-2 strain that is now dominant worldwide but was not prevalent in earlier vaccine studies.

## Limitations

This study has several limitations. First, given the observational design, there is potential for unmeasured confounding factors. In particular, patients included in this study were vaccinated individuals who chose to request a RT-PCR test for SARS-CoV-2 during the study period. Individuals may have variable thresholds for requesting a test and may sometimes do so for reasons other than disease related complaints. Despite this limitation, given the high magnitude of the observed risk difference, it is unlikely that unmeasured bias could account to the full extent of the risk increase observed in patients who had an older vaccine.

Since most of the new infections were observed in the last two weeks, and given the lag usually observed between the onset of symptoms and the need for hospitalization, it is too early to assess the severity of these new infections in terms of need for hospital admission, need for mechanical ventilation or mortality. These data should be followed carefully to assess whether the increased risk infection rate seen in individuals with an older vaccine is also associated with increased risks of severe complications.

It is also not known how long any protective effect of mRNA vaccination and SARS-CoV-2 infection may last beyond the studied days in either group. These questions remain to be addressed by further research.

## Conclusions

In this retrospective large cohort study performed on patients fully vaccinated with BNT162b2 mRNA vaccine, protection appeared to decrease over time, with risk for SARS-CoV-2 infection significantly higher in patients who received their second vaccine dose at least 146 days ago. Interpretation of study findings is limited by the observational design; but may warrant the consideration of an additional vaccine dose in individuals at risk for severe disease.

## Data Availability

Data were obtained from patients' electronic health records, and IRB approval restrains its use to researchers inside Leumit Health Services.

## Article Information

Dr. Israel had full access to all of the data in the study and takes responsibility for the integrity of the data and the accuracy of the data analysis.

## Conflict of Interest Disclosures

None reported.

## Acknowledgment

This research was supported in part by the Intramural Research Program of the National Institutes of Health, National Cancer Institute.

## Notes

### Competing Interest Statement

The authors have declared no competing interest.

### Author Declarations

the Shamir Medical Center Institutional Review Board (129-2-LEU).

